# Association between metabolic syndrome and humoral immune response to Pfizer–BioNTech vaccine in healthcare workers

**DOI:** 10.1101/2022.04.13.22273853

**Authors:** Dong Van Hoang, Shohei Yamamoto, Ami Fukunaga, Yosuke Inoue, Tetsuya Mizoue, Norio Ohmagari

## Abstract

**Purpose:** The clustering of metabolic abnormalities may weaken vaccine-induced immunity, but epidemiological data regarding SARS-CoV-2 vaccines are scarce. The present study examined the cross-sectional association between metabolic syndrome (MetS) and humoral immune response to Pfizer–BioNTech vaccine among healthcare workers.

**Methods:** Participants were 946 healthcare workers, aged 21–75 years, who had completed the second dose of Pfizer–BioNTech vaccine 1–3 months before the survey. MetS was defined according to the Joint Interim Statement. SARS-CoV-2 spike immunoglobulin G (IgG) antibody was measured using quantitative assays. Multivariable linear regression was used to estimate the geometric mean titers (GMT) and geometric mean ratio (GMR) of IgG titers, relative to MetS status.

**Results:** A total of 51 participants (5.4%) had MetS. Healthcare workers with MetS had a significantly lower IgG titer (GMT 3882; 95% confidence interval [CI], 3124–4824) than those without MetS (GMT 5033; 95% CI, 4395–5764); the GMR was 0.77 (95% CI 0.64–0.93). The GMR for IgG titers among those having 0 (reference group), 1, 2, 3, or ≥ 4 MetS components was 1.00, 1.00, 0.89, 0.86 and 0.61, respectively (P_trend_ = 0.004).

**Conclusion:** Results suggest that having MetS and a greater number of its components are associated with a weaker humoral immune response to the Pfizer–BioNTech vaccine.

## 1. Introduction

The ongoing pandemic of severe acute respiratory syndrome coronavirus (SARS-CoV-2) has infected more than 450 million people, caused more than 6 million deaths globally, and continues to spread worldwide [1]. Immunization with SARS-CoV-2 vaccines can be a global strategy to minimize deaths, severity, and overall disease burden of the pandemic [2]. While most recommended SARS-CoV-2 vaccines, such as BioNTech (BNT162b2) and Moderna (mRNA-1273) can achieve high efficacy [3], their immunogenicity can be hampered by several factors, e.g., aging, virus mutation [4,5], smoking [5], obesity [6,7], diabetes [7], and other underlying comorbidities [5]. Identification of such factors may be of public health significance regarding the prevention of the virus infection, e.g., administration of an earlier vaccine booster in high-risk groups [8].

Epidemiological data suggest that metabolic syndrome (MetS), a major public health concern for many countries worldwide [9,10], may hurt the humoral response to SARS-CoV-2 vaccines. More specifically, MetS can lead to a chronic inflammatory state (e.g., increased circulating adipokines and cytokine-like hormones) which in turn may result in a decrease in immunogenicity following vaccination [11–13]. A few studies [7,14,15] showed that individual MetS components may reduce the immune response to SARS-CoV-2 vaccines. For example, central obesity was associated with lower immunoglobulin (Ig) G antibody titers [14]; while diabetes was inversely associated with IgG antibody concentration [15], after the vaccination with Pfizer–BioNTech. However, we are not aware of any epidemiological data linking MetS to immune response to SARS-CoV-2 vaccines. Here, we examined SARS-CoV-2 spike IgG antibody titers in relation to MetS among vaccine recipients.

## 2. Method

### 2.1. Study setting and participants

We are conducting a repeatitive serological survey among the staff members of the National Center for Global Health and Medicine (NCGM, consisting of Toyama and Kohnodai hospitals), Japan to monitor the spread of SARS-CoV-2 infection, with the first round launched in July 2020 [16–18]. We collected the information on medical history, health-related lifestyle, COVID-19 related information (e.g., COVID-19 infection and vaccination), and a blood sample to measure the participants’ serum levels of SARS-CoV-2 nucleocapsid and spike-protein antibodies. We additionally obtained information collected during annual health check-up which was conducted in the same year as the survey (October 2020 and June 2021). Written informed consent was obtained from each participant, and the study procedure was approved by the ethics committee of NCGM (NCGM-G-003598).

The present study used data of the third survey in June 2021, 2 months after the completion of an in-house vaccination program (Pfizer–BioNTech). Of 3,072 NCGM’s workers invited, 2,779 (90%) participated. Among the participants, 2479 had received two doses of Pfizer–BioNTech vaccine, but we excluded those who did not agree to provide their health check-up data (n = 202) and those who attended the survey within 14 days of their second vaccination (n = 5). Since the information on the fasting status at the time of blood testing was not available for participants from Kohnodai hospital (n = 472), we only included participants from Toyama hospital (n = 1,800). We further excluded those with missing information on waist circumference (WC) (n = 266), or those with non-fasting plasma glucose (n = 588), leaving an analytical sample of 946 participants. Those who were excluded (n = 1533) were older, more likely to be women, smoked more, and had a higher prevalence of comorbidities and history of SARS-CoV-2 infection, and higher median of SARS-CoV-2 spike IgG titers (Supplementary table 1).

**Table 1:** Characteristics of study participants.

| Characteristics | All participants | Metabolic syndrome |  |
| --- | --- | --- | --- |
|  |  | No | Yes |
| N | 946 | 895 | 51 |
| Age (year), mean [SD] | 36.7 [12.3] | 35.9 [12.0] | 49.5 [10.7] |
| Sex (men) | 298 (31.5) | 271 (30.3) | 27 (52.9) |
| Smoking |  |  |  |
| Non-smoker | 855 (90.4) | 819 (91.5) | 36 (70.6) |
| Smoker | 91 (9.6) | 76 (8.5) | 15 (29.4) |
| Occupation |  |  |  |
| Nurse | 285 (30.1) | 279 (31.2) | 6 (11.8) |
| Doctor | 183 (19.3) | 171 (19.0) | 12 (23.5) |
| Administrative staff | 134 (14.2) | 125 (14.0) | 9 (17.6) |
| Allied healthcare professionals | 125 (13.2) | 119 (13.3) | 6 (11.8) |
| Others | 219 (23.2) | 201 (22.5) | 18 (35.3) |
| Alcohol consumption |  |  |  |
| Non-drinker | 355 (37.5) | 332 (37.1) | 23 (45.1) |
| Drinker consuming |  |  |  |
| < 23 g ethanol/day | 442 (46.7) | 424 (47.4) | 18 (35.3) |
| ≥ 23 g ethanol/day | 149 (15.8) | 139 (15.5) | 10 (19.6) |
| Leisure time physical activity |  |  |  |
| Non-engagement | 197 (20.8) | 185 (20.7) | 12 (23.5) |
| < 150 min/week | 657 (69.5) | 625 (69.8) | 32 (62.7) |
| ≥ 150 min/week | 92 (9.7) | 85 (9.5) | 7 (13.7) |
| Comorbidities | 29 (3.1) | 25 (2.8) | 4 (7.8) |
| Lung disease | 18 (1.9) | 16 (1.8) | 2 (3.9) |
| Heart disease | 5 (0.5) | 4 (0.4) | 1 (2.0) |
| Cancer | 6 (0.6) | 5 (0.6) | 1 (2.0) |
| History of SARS-Cov-2 infection <sup>a</sup> | 5 (0.5) | 5 (0.6) | 0 (0.0) |
| Vaccine-to-IgG test days, median [range] <sup>b</sup> | 67 [15-103] | 67 [15-103] | 69 [35-98] |
| SARS-Cov-2 spike IgG (AU/mL), GMT [SD] | 5415 [2.2] | 5606 [2.1] | 2947 [2.6] |

Values are n (%), unless otherwise stated; AU: antibody unit; GMT: geometric mean titers; SD: standard deviation.
<sup>a</sup> defined as the positive result of either polymerase chain reaction test or the measurement of antibodies against SARS-CoV-2 nucleocapsid protein.
<sup>b</sup> time interval between the second dose of vaccine and the day of blood draw

### 2.2. Assessment of SARS-CoV-2 antibodies

We quantitatively measured IgG (AU/mL) against the SARS-CoV-2 spike protein, using AdviseDx SARS-CoV-2 IgG II assay, Abbott ARCHITECT^®^. In a subgroup of vaccine recipients in this cohort, the SARS-CoV-2 spike IgG titers measured with this assay had a strong correlation with neutralizing antibody titers (Spearman’s ρ=0.91) [19]. We also qualitatively measured antibodies against SARS-CoV-2 nucleocapsid protein using the SARS-CoV-2 IgG assay (Abbott) and used these data to identify those with possible infection in the past. The sensitivity and specificity for the identification of past infection with SARS-CoV-2 viruses using this assay were 100% and 99.9%, respectively [20].

### 2.3. Assessment of metabolic syndrome and covariates

The information on MetS components, i.e., WC, fasting plasma glucose (FPG), blood pressure (BP), triglycerides (TG), and high-density lipoprotein cholesterol (HDL-C) was collected during the health check-up. MetS was defined, according to the Joint Interim Statement [21], as clustering of any three or more of the following components: high FPG (≥ 100 mg/dL or using anti-diabetic medication), central obesity (WC ≥ 90 cm for men, or ≥ 80 cm for women), high TG (≥ 150 mg/dL or using lipid-lowering medication), high BP (systolic BP ≥ 130 mmHg, diastolic BP ≥ 85 mmHg or using antihypertensive medication) and reduced HDL-C (< 40 mg/dL for men or < 50 mg/dL for women). The cut-off values for WC were based on the recommendation of the World Health Organization for Asian populations [22].

We selected covariates according to epidemiological evidence for their association with the immune response to SARS-CoV-2 vaccines: age, sex [4,5], smoking [5], alcohol drinking [23], physical activity [24], underlying comorbidities (i.e., cancer, heart, or lung diseases) [5,25], history of SARS-CoV-2 infection [23,26], and the time interval between the second dose of SARS-CoV-2 vaccine and the day of blood draw (vaccine-to-IgG test days) [23]. The history of infection with SARS-CoV-2 was defined as the positive result of either polymerase chain reaction test or antibodies against SARS-CoV-2 nucleocapsid protein.

### 2.4 Statistical analysis

The background characteristics of the study population, according to MetS status, were described as arithmetic/geometric mean and standard deviation (SD), or median and range for continuous variables, and percentages for categorical variables.

Linear regression modeling was used to estimate the means (95% confidence interval [CI]), and the beta-coefficients (95% CI) of log_10_-transformed SARS-CoV-2 spike IgG titers, relative to MetS. Two models were fitted: Model 1 was adjusted for age and sex; and Model 2 was further adjusted for smoking (non-smoker, or smoker), alcohol drinking (non-drinker, drinker consuming < 23 or ≥ 23 g ethanol/day), leisure-time physical activity (non-engagement, < 150, or ≥ 150 min/week), comorbidity of cancer, heart or lung diseases, history of SARS-CoV-2 infection, and vaccine-to-IgG test days. The estimated values were then back-transformed to obtain the geometric mean titers (GMT) (95% CI) and geometric mean ratio (GMR) (95% CI) of SARS-CoV-2 spike IgG. We also examined the association between the number of MetS components and SARS-CoV-2 spike IgG titers, using Model 1 and Model 2 in which those with five components were regrouped together with those having four components. The trend in this association was assessed by assigning an ordinal number (1 to 5) to each group, which was treated as a continuous variable when fitted in regression models.

To eliminate the potential impact of comorbidities and history of SARS-CoV-2 infection on the association between MetS and the immunogenicity of Pfizer–BioNTech vaccine, we conducted a sensitivity analysis using Model 1 and Model 2 in which we excluded participants with either condition. Statistical significance was set at p < 0.05 for trend and p < 0.1 for interaction tests. All statistical analyses were conducted in RStudio (version 3.2.4) using the package “emmeans” (version 1.6.3) [27].

## 3. Results

**Table 1** presents the characteristics of study participants. Workers with MetS accounted for 5.4% of the study participants, were older, more likely to be men, smoked and drank more, and had a higher prevalence of comorbidities compared with those without the syndrome. They also engaged more in leisure-time physical activity. In contrast, they had lower GMT of SARS-CoV-2 spike IgG antibody titers.

**Table 2** presents the associations of MetS and the number of MetS components with SARS-CoV-2 spike IgG titers. Participants with MetS showed a significantly lower GMT of SARS-CoV-2 spike IgG, compared with those without MetS; the age and sex-adjusted GMR (95% CI) was 0.77 (0.63-0.94). After further adjusting for smoking, alcohol consumption, physical activity, comorbidity of cancer, heart, or lung diseases, history of SARS-CoV-2 infection, and vaccine-to-IgG test days, the association remained almost unchanged (GMR 0.77; 95% CI, 0.64–0.93). SARS-CoV-2 spike IgG titers steadily decreased with increasing number of MetS components; the GMR for those having 0 (reference group), 1, 2, 3 or ≥ 4 components was 1.00, 1.00, 0.89, 0.86 and 0.61, respectively (P_trend_ = 0.004). In the sensitivity analysis excluding those with a history of SARS-CoV-2 infection or those with comorbid cancer, heart diseases, or lung diseases (Supplementary table 2), the results were virtually unchanged.

**Table 2:** Association between MetS and SARS-Cov-2 spike IgG titers.

| MetS/<br>number of<br>MetS<br>components | N | SARS-Cov-2 spike IgG |  |  |  |
| --- | --- | --- | --- | --- | --- |
|  |  | Model 1 |  | Model 2 |  |
|  |  | GMT (95% CI) | GMR (95% CI) | GMT (95% CI) | GMR (95% CI) |
| Metabolic syndrome |  |  |  |  |  |
| MetS (-) | 895 | 5268 (5012, 5538) | 1.00 (ref) | 5033 (4395, 5764) | 1.00 (ref) |
| MetS (+) | 51 | 4053 (3322, 4944) | 0.77 (0.63, 0.94) | 3882 (3124, 4824) | 0.77 (0.64, 0.93) |
| Number of metabolic syndrome components |  |  |  |  |  |
| 0 | 572 | 5375 (5038, 5735) | 1.00 (ref) | 5090 (4421, 5860) | 1.00 (ref) |
| 1 | 233 | 5318 (4850, 5832) | 0.99 (0.88, 1.11) | 5108 (4390, 5944) | 1.00 (0.90, 1.11) |
| 2 | 90 | 4644 (3994, 5400) | 0.86 (0.73, 1.02) | 4553 (3751, 5525) | 0.89 (0.77, 1.04) |
| 3 | 32 | 4333 (3376, 5560) | 0.81 (0.62, 1.05) | 4380 (3377, 5682) | 0.86 (0.68, 1.10) |
| ≥ 4 | 19 | 3488 (2535, 4800) | 0.65 (0.47, 0.90) | 3089 (2257, 4227) | 0.61 (0.45, 0.82) |
| P for trend |  |  | 0.004 |  | 0.004 |
| MetS: Metabolic syndrome; GMT: geometric mean titers; GMR: geometric mean ratio; ref: reference; Model 1: adjusted for age and sex; Model 2: adjusted for age, sex, smoking, alcohol consumption, leisure time physical activity, comorbidity of cancer, cardiovascular or lung diseases, time interval (in day) between the second vaccination and the day of blood draw, and History of SARS-Cov-2 infection. |  |  |  |  |  |

## 4. Discussion

In the present cross-sectional study, MetS was associated with a significantly lower SARS-CoV-2 spike IgG antibody titer among healthcare workers who received two doses of Pfizer– BioNTech vaccine. There was also a significant inverse association between the number of MetS components and SARS-CoV-2 spike IgG antibody titer.

We are not aware of any previous studies on the relationship between MetS and SARS-CoV-2 spike IgG antibody titers. Nevertheless, our finding is in line with epidemiological data on the association between individual MetS components and the immunogenicity of SARS-CoV-2 vaccines. For example, in Italian healthcare workers, central obesity (p = 0.026) [14], overweight (p = 0.04) [28], and dyslipidemia (p = 0.005) were each associated with lower IgG antibody titers following Pfizer–BioNTech vaccination. A systematic review of eight studies showed that diabetic patients had weaker immunogenicity following SARS-CoV-2 vaccines (i.e., Pfizer–BioNTech, CoronaVac, and Covishield™), compared with healthy people [15]. In the present study, we also observed an inverse dose-response association between the number of MetS components and Pfizer–BioNTech-induced antibody titers, which further consolidated our observed association between MetS and the weaker immune response to this vaccine.

One possible explanation for the present association could be due to the MetS-induced chronic systemic inflammation. In the condition of MetS, several pro-inflammatory cytokines (e.g., leptin, TNF-α, and IL-6) are over-secreted, while some anti-inflammatory cytokines (e.g., adiponectin) are under-secreted [12]. Subsequently, this dysregulated production of adipokines results in chronic low-grade inflammation, which in turn may lead to an alteration in the function of B cells, and a subsequent reduction in vaccine-induced antibody production [11,12,29,30].

This study has some limitations. First, we excluded a large number of participants who lacked biochemical data obtained in the fasting state, leading to a decrease in the precision of estimates due to the reduced sample size. Second, immunosuppressive medication may confound the association between MetS and vaccine immunogenicity. However, we did not obtain information about the use of such medication in the third survey. Third, the findings were derived from healthy healthcare workers, and thus, the generalization should be made with caution.

In conclusion, this study among healthcare workers showed that MetS was associated with significantly lower concentrations of SARS-CoV-2 spike IgG antibody titers. Further studies should clarify whether individuals with MetS have a higher risk of COVID-19 breakthrough infection and thus are recommended to receive vaccines in a short interval of time.

## Data Availability

The datasets generated and/or analyzed during the current study are not publicly available due to ethical restrictions and participant confidentiality concerns.

## Abbreviations^1^

^1^MetS: metabolic syndrome
Ig: immunoglobulin
GMT: geometric mean titers
GMR: geometric mean ratio
CI: confidence interval
SARS-CoV-2: severe acute respiratory syndrome coronavirus
NCGM: National Center for Global Health and Medicine
FPG: fasting plasma glucose
BP: blood pressure
TG: triglycerides
HDL-C: high-density lipoprotein cholesterol

## Contributors

DVH performed the data analyses and manuscript drafting; SY, AF, conducted data acquisition; AF, YI, TM, and NO were involved in the interpretation of the results and revision of the manuscript. D.V.H. and TM took primary responsibility for the final content. All authors have read and agreed to the published version of the manuscript.

## Acknowledgements

We thank Haruka Osawa for her contribution to data collection.

## Data Availability

The datasets generated and/or analyzed during the current study are not publicly available due to ethical restrictions and participant confidentiality concerns, but de-identified data are available from the corresponding author to qualified researchers on reasonable request.

## Funding

This work was supported by the NCGM COVID-19 Gift Fund (grant number 19K059), and the Japan Health Research Promotion Bureau Research Fund (grant number 2020-B-09).

## Declaration of competing interest

Antibody assay reagent was provided by Abbott Japan.

## Notes

### Competing Interest Statement

The authors have declared no competing interest.

### Author Declarations

The ethics committee of National Center for Global Health and Medicine

