## Supplemental tables for "Association between metabolic syndrome and humoral immune response to Pfizer–BioNTech vaccine in healthcare workers"

### Supplementary table 1: Characteristics of included versus excluded participants

| Characteristics | Excluded participants | Included participants |
|---|---|---|
| N | 1753 | 946 |
| Age, mean [SD] | 38.9 (12.0) | 36.7 (12.3) |
| Sex (men) | 484 (27.6) | 298 (31.5) |
| Smoking | | |
| Non-smoker | 1482 (84.5) | 855 (90.4) |
| Smoker | 271 (15.5) | 91 (9.6) |
| Alcohol consumption | | |
| Non-drinker | 713 (40.7) | 355 (37.5) |
| Drinker consuming | | |
| < 1 go/day | 757 (43.3) | 442 (46.7) |
| ≥ 1 go/day | 280 (16.0) | 149 (15.8) |
| Missing | 3 | |
| Leisure time physical activity | | |
| Non-engagement | 375 (21.4) | 197 (20.8) |
| < 150 min/week | 1220 (69.7) | 657 (69.5) |
| ≥ 150 min/week | 156 (8.9) | 92 (9.7) |
| Missing | 2 | |
| Comorbidity | 91 (5.2) | 29 (3.1) |
| Lung disease | 62 (3.5) | 18 (1.9) |
| Heart disease | 22 (1.3) | 5 (0.5) |
| Cancer | 17 (1.0) | 6 (0.6) |
| History of SARS-Cov-2 infection [a] | 27 (1.5) | 5 (0.5) |
| Vaccine-to-IgG test days, median [range] [b] | 62 [5-103] | 67 [15-103] |
| Missing | 220 | 0 |
| SARS-Cov-2 spike IgG (AU/mL), median [range] | 5708 [0-61922] | 5588 [135-36962] |
| Missing | 16 | 0 |
| Central obesity [c] | 203 (21.9) | 187 (19.8) |
| Missing | 826 | |
| High BP [d] | 256 (25.1) | 213 (22.5) |
| Missing | 732 | |
| High FPG [e] | 17 (18.3) | 65 (6.9) |
| Missing | 1660 | |
| High TG [f] | 159 (16.0) | 93 (9.8) |
| Missing | 761 | |
| Reduced HDL-C [g] | 45 (3.7) | 30 (3.2) |
| Missing | 535 | |

Values are n (%), unless otherwise stated; SD: standard deviation; IgG: immunoglobulin G;
[a] positive result of either polymerase chain reaction test or the measurement of antibodies against SARS-CoV-2 nucleocapsid protein; [b] time interval between the second dose of vaccine and the day of blood draw;
[c] waist circumference ≥ 90 cm for men, or ≥ 80 cm for women; [d] systolic blood pressure [BP] ≥ 130 mmHg, diastolic BP ≥ 85 mmHg or using antihypertensive medication; [e] fasting plasma glucose ≥ 100 mg/dL or using antidiabetic medication; [f] triglycerides ≥ 150 mg/dL or using lipid-lowering medication; [g] high-density lipoprotein cholesterol < 40 mg/dL for men, or < 50 mg/dL for women.

**Supplementary table 2: Association between MetS and SARS-Cov-2 spike IgG titers, excluding those with history of SARS-Cov-2 infection and those with comorbid cancer, heart or lung diseases**

| MetS/ number of MetS components | N | SARS-Cov-2 spike IgG | | | |
|---|---|---|---|---|---|
| | | Model 1 | | Model 2 | |
| | | GMT (95% CI) | GMR (95% CI) | GMT (95% CI) | GMR (95% CI) |
| | | **Metabolic syndrome** | | | |
| MetS (-) | 865 | 5309 (5049, 5582) | 1.00 (ref) | 5051 (4636, 5503) | 1.00 (ref) |
| MetS (+) | 47 | 4020 (3278, 4929) | 0.76 (0.61, 0.93) | 3816 (3136, 4644) | 0.76 (0.62, 0.92) |
| | | **Number of metabolic syndrome components** | | | |
| 0 | 552 | 5387 (5046, 5750) | 1.00 (ref) | 5057 (4593, 5568) | 1.00 (ref) |
| 1 | 225 | 5425 (4944, 5952) | 1.01 (0.90, 1.13) | 5217 (4686, 5807) | 1.03 (0.93, 1.15) |
| 2 | 88 | 4678 (4022, 5440) | 0.87 (0.73, 1.03) | 4550 (3882, 5331) | 0.90 (0.77, 1.05) |
| 3 | 30 | 4233 (3282, 5459) | 0.79 (0.60, 1.03) | 4245 (3335, 5404) | 0.84 (0.66, 1.07) |
| ≥ 4 | 17 | 3554 (2548, 4957) | 0.66 (0.47, 0.93) | 3094 (2270, 4218) | 0.61 (0.45, 0.84) |
| P for trend | | | 0.006 | | 0.006 |

MetS: Metabolic syndrome; GMT: geometric mean titers; GMR: geometric mean ratio; ref: reference; Model 1: adjusted for age and sex; Model 2: adjusted for age, sex, smoking, alcohol consumption, leisure time physical activity, and time interval (in day) between the second vaccination and the day of blood draw
